# Prevalence of Anemia and Associated Factors Among Post-Partum Mothers in Public Health Facilities in Ethiopia: A Systematic Review and Meta-Analysis

**DOI:** 10.64898/2026.07.12.26357855

**Authors:** Hermella Negash Woldearegay, Maedot Sebsibe Tebeka, Saeed Omer Saeed Osman

## Abstract

**Background:** Postpartum anemia (PPA) is defined as a hemoglobin concentration below 11 g/dL within the first week following delivery, or below 12 g/dL at eight weeks postpartum. It constitutes a major yet under-addressed public health burden in sub-Saharan Africa, contributing substantially to maternal morbidity and mortality. In Ethiopia, anemia prevalence among women of reproductive age has been increasing despite national nutrition strategies, yet aggregated, nationally representative data on PPA remain scarce. This systematic review and meta-analysis aimed to estimate the pooled prevalence of anemia among postpartum mothers attending public health facilities in Ethiopia and to identify significantly associated factors.

**Methods:** A comprehensive literature search was conducted across PubMed/MEDLINE, Cochrane Library, Google Scholar, African Journals Online (AJOL), and HINARI from inception to December 2024. The Preferred Reporting Items for Systematic Reviews and Meta-Analyses (PRISMA 2020) guidelines were followed. Observational studies (cross-sectional, cohort) conducted in public health facilities in Ethiopia, reporting hemoglobin-confirmed PPA prevalence and/or associated factors, were included. Two reviewers independently performed study selection, data extraction, and quality assessment using a modified Newcastle-Ottawa Scale (NOS) adapted for cross-sectional studies. Heterogeneity was quantified using the Cochran Q statistic and I-squared index. Pooled prevalence estimates and odds ratios were computed using a random-effects (DerSimonian-Laird) model with 95% confidence intervals. Subgroup analyses were performed by geographic region (Eastern vs. Western/Central Ethiopia), study period, and facility type. Publication bias was assessed via Egger weighted regression test and funnel plot asymmetry.

**Results:** Six primary studies, encompassing 2,819 postpartum mothers, fulfilled the eligibility criteria and were included in the meta-analysis. The pooled prevalence of anemia among postpartum mothers in Ethiopian public health facilities was 35.4% (95% CI: 27.6-43.6%; I2 = 89.2%), classified as a severe public health problem per WHO thresholds. Subgroup analysis revealed higher prevalence in Eastern Ethiopia (Dire Dawa and Harari regions: 27.5%) compared to Western/Central Ethiopia (Gondar and Debre Markos: 24.3-47.1%), though marked heterogeneity was observed. Factors significantly associated with higher odds of PPA included: fewer than four antenatal care (ANC) visits (pooled OR = 2.72; 95% CI: 2.14-3.30), history of postpartum hemorrhage (OR = 2.49; 95% CI: 1.08-3.98), cesarean section delivery (OR = 4.04; 95% CI: 3.43-4.67), instrumental delivery forceps/vacuum (OR = 3.96; 95% CI: 2.99-4.95), poor adherence to iron and folic acid (IFA) supplementation (OR = 2.80; 95% CI: 2.31-3.30), low pre-delivery hemoglobin < 11 g/dL (OR = 4.20; 95% CI: 1.77-6.67), low dietary diversity (OR = 4.20; 95% CI: 1.77-6.67), and lack of formal education (OR = 3.50; 95% CI: 2.64-4.41).

**Conclusions:** Nearly one in three postpartum mothers attending public health facilities in Ethiopia is anemic, constituting a severe public health emergency. The burden is driven by modifiable clinical and behavioral factors — particularly inadequate ANC utilization, poor IFA adherence, hemorrhagic complications, and nutritional deficiencies — as well as structural determinants including low educational attainment. Evidence-based, multi-sectoral interventions are urgently needed, including strengthening ANC quality and coverage, universal IFA supplementation monitoring, active postpartum hemorrhage management, and targeted nutritional counseling. Policy frameworks must address regional disparities and integrate anemia screening into routine postpartum care protocols.

## 1. Background

Anemia is defined by the World Health Organization (WHO) as a hemoglobin (Hb) concentration below established sex- and physiological state-specific thresholds that remains one of the most prevalent nutritional disorders worldwide. In postpartum women, the diagnostic threshold is set at Hb < 11 g/dL (or hematocrit < 33%) within the first week after delivery, and Hb < 12 g/dL at eight weeks postpartum, distinguishing it from both pregnancy-related and non-pregnancy-related anemia [1,2]. The postpartum period represents a particularly vulnerable window: physiological hemodynamic shifts, obligate blood loss at delivery, lactational iron demands, and the cumulative toll of antepartum nutritional deficiencies all converge to elevate anemia risk substantially.

Globally, anemia affected an estimated 613 million (33%) of non-pregnant women of reproductive age in 2016, with the highest burdens concentrated in South Asia and sub-Saharan Africa [3]. In high-income countries, PPA prevalence ranges from approximately 10% to 30%, while in low- and middle-income countries (LMICs) particularly in sub-Saharan Africa and South Asia prevalence is reported between 50% and 80% [4,5]. The consequences of PPA extend beyond the mother: impaired cognitive function, emotional instability, heightened risk of postpartum depression, reduced breast milk quality and quantity, and at severe levels direct cardiovascular compromise and maternal mortality [6,7].

Ethiopia represents a critical case study in the epidemiology of postpartum anemia. The 2016 Ethiopian Demographic and Health Survey (EDHS) documented anemia prevalence of 28.6% among lactating mothers a significant increase from 18.6% in the 2011 survey despite the country’s National Nutrition Program (NNP) targeting a reduction to 12% by 2020 [8,9]. Regional heterogeneity is substantial, with studies from the Amhara region (Debre Markos) reporting 24.3% prevalence, while studies from Eastern Ethiopia (Dire Dawa and Harari) document prevalences of 26.9% and 28.1%, respectively [10,11]. A 2023 facility-based study in Gondar, Northwest Ethiopia, found PPA as high as 47.1% [12].

Despite growing recognition of PPA as a public health problem in Ethiopia, existing individual studies are limited by small sample sizes, geographic specificity, and varied methodological approaches making national inference difficult. Prior systematic reviews have either focused narrowly on the immediate postpartum period, confined analysis to specific sub-populations, or included only a limited number of primary studies, leaving considerable uncertainty in pooled prevalence estimates and in the relative importance of associated factors. A methodologically rigorous, comprehensive systematic review and meta-analysis synthesizing all available evidence from across Ethiopia is therefore urgently needed to inform policy and programmatic responses.

This study aimed to:

(i) estimate the pooled prevalence of anemia among postpartum mothers attending public health facilities in Ethiopia;
(ii) identify and quantify factors independently associated with PPA.
(iii) examine subgroup differences by geographic region and study period, to provide actionable evidence for health policy formulation and targeted interventions.

## 2. Methods

This systematic review and meta-analysis were conducted and reported in accordance with the PRISMA 2020 guidelines [13].

### 2.1 Eligibility Criteria

Studies were included if they met the following criteria (PICOS framework):

- **Population:** Postpartum mothers (women within 6 weeks of delivery) aged ≥ 15 years attending public health facilities in Ethiopia, regardless of parity or mode of delivery.
- **Intervention/Exposure:** Not applicable (observational studies). Exposure variables included individual, obstetric, clinical, and socioeconomic factors.
- **Comparator:** Non-anemic postpartum mothers (for factor analysis); general population benchmarks (for prevalence estimates).
- **Outcomes:** Primary: Prevalence of anemia (Hb < 11 g/dL confirmed by laboratory hematological analysis). Secondary: Adjusted odds ratios (AOR) for associated factors.
- **Study Design:** Cross-sectional, cohort (prospective or retrospective), and case-control studies published in peer-reviewed journals or as grey literature.
- **Setting:** Public health facilities (health centers, district hospitals, referral hospitals) in any region of Ethiopia.
- **Language & Time:** No language restriction. Studies published from January 2010 to December 2024.

**Exclusion criteria**

- Studies conducted in private facilities
- Studies using self-reported or clinical (non-laboratory) anemia diagnosis
- Conference abstracts without retrievable full texts; duplicate publications from the same study population.

### 2.2 Information Sources and Search Strategy

A systematic search was conducted across the following databases:

PubMed/MEDLINE, Cochrane Central Register of Controlled Trials, Google Scholar, African Journals Online (AJOL), HINARI, and WHO IRIS. Grey literature was sought via OpenDOAR, Addis Ababa University Institutional Repository, and direct author contact. The search was last updated on 31 July 2024.

The search strategy used the following MeSH terms and free-text keywords in Boolean combination:

> *(“Anemia” OR “Anaemia” OR “low hemoglobin” OR “iron deficiency”) AND (“postpartum” OR “postnatal” OR “after delivery” OR “puerperal”) AND (“prevalence” OR “magnitude” OR “proportion”) AND (“Ethiopia” OR “Ethiopian” OR “Amhara” OR “Oromia” OR “SNNPR” OR “Tigray” OR “Afar” OR “Somali” OR “Harari” OR “Dire Dawa” OR “Addis Ababa”)*

The complete PubMed search string is provided in Supplementary File S1. Forward and backward citation tracking was performed on all included studies.

### 2.3 Study Selection

All retrieved records were imported into Rayyan systematic review software for duplicate removal and title/abstract screening.

Two reviewers (Author 1 and Author 2) independently screened all records in two sequential phases:

(i) title and abstract screening against predefined eligibility criteria and
(ii) full-text assessment of potentially eligible studies.

Disagreements were resolved by consensus or arbitration by a third reviewer (Author 3).

### 2.4 Data Extraction

A standardized data extraction form as stipulated under Annex 2 was piloted on five studies and refined prior to full implementation. Two reviewers independently extracted the following information:

(1) study identification (authors, year, journal, DOI)
(2) study design, setting, and geographic region;
(3) sampling method and sample size;
(4) participant characteristics (age, parity, education)
(5) anemia diagnostic criteria and laboratory method
(6) anemia prevalence and 95% CI
(7) associated factors reported as OR/AOR with 95% CI;
(8) confounders adjusted for and
(9) quality score.

Discrepancies were adjudicated by consensus.

### 2.5 Quality Assessment

Methodological quality of included studies was evaluated using the Newcastle-Ottawa Scale (NOS) adapted for cross-sectional studies [14].

The adapted tool assesses three domains:

(A) **Selection (4 items, maximum 4 stars):** Representativeness of the sample, sample size justification, non-respondents’ description, and ascertainment of exposure/outcome using validated instrument;
(B) **Comparability (2 items, maximum 2 stars):** Whether studies controlled for the most important confounders (education, parity, ANC visits);
(C) **Outcome (3 items, maximum 3 stars):** Assessment of outcome by laboratory hemoglobin measurement, appropriate follow-up, and statistical adequacy.

Studies scoring ≥ 7 stars were classified as high quality, 4–6 as moderate, and ≤ 3 as low quality. Low-quality studies were included in sensitivity analyses to assess their influence on pooled estimates.

### 2.6 Statistical Analysis

All statistical analyses were performed using R statistical software (version 4.4.0) with the “meta” (v6.5-0) and “metafor” (v4.4-0) packages. The following analytical approach was used:

1. Prevalence estimates from individual studies were transformed using the double-arcsine (Freeman–Tukey) transformation to stabilize variance prior to meta-analysis, then back-transformed for reporting.
2. Pooled prevalence and pooled odds ratios were estimated using the DerSimonian-Laird random-effects model, which accounts for between-study heterogeneity.
3. Heterogeneity was quantified using:

(i) the Cochran Q test (significance threshold: p < 0.10)
(ii) the I² statistic (thresholds: < 25% low, 25–75% moderate, > 75% high) and
(iii) the τ² (Tau-squared) estimate of between-study variance.
4. Subgroup analyses were pre-specified by: geographic region (Eastern vs. Western/Central Ethiopia), study period (pre-2020 vs. 2020 onwards), facility type (health center vs. hospital), and NOS quality category (high vs. moderate quality).
5. Meta-regression was performed to explore potential sources of heterogeneity, with covariates including year of publication, sample size, and regional location.
6. Publication bias was assessed by visual inspection of funnel plots and formally tested using Egger’s weighted regression test (significance threshold: p < 0.10). Trim-and-fill analysis (Duval and Tweedie method) was applied if asymmetry was detected.
7. Sensitivity analyses were conducted by sequentially omitting one study at a time (leave-one-out analysis) and by restricting the analysis to high-quality studies (NOS ≥ 7).

All tests were two-tailed. A p-value < 0.05 was considered statistically significant unless otherwise specified.

## 3. Results

### 3.1 Study Selection

The systematic database search yielded 847 records (PubMed: 312; Cochrane Library: 89; Google Scholar: 318; AJOL/HINARI: 128). Following removal of 234 duplicates, 613 records underwent title and abstract screening. Of these, 581 were excluded based on predefined criteria. Thirty-two full-text articles were assessed for eligibility; 26 were subsequently excluded (reasons detailed in PRISMA flow diagram, Figure 1). Six studies met all inclusion criteria and were incorporated into the meta-analysis, encompassing a combined sample of 2,819 postpartum mothers. Inter-rater agreement was excellent at both the title/abstract screening phase (κ = 0.84) and the full-text phase (κ = 0.91).

**FIGURE 1.**
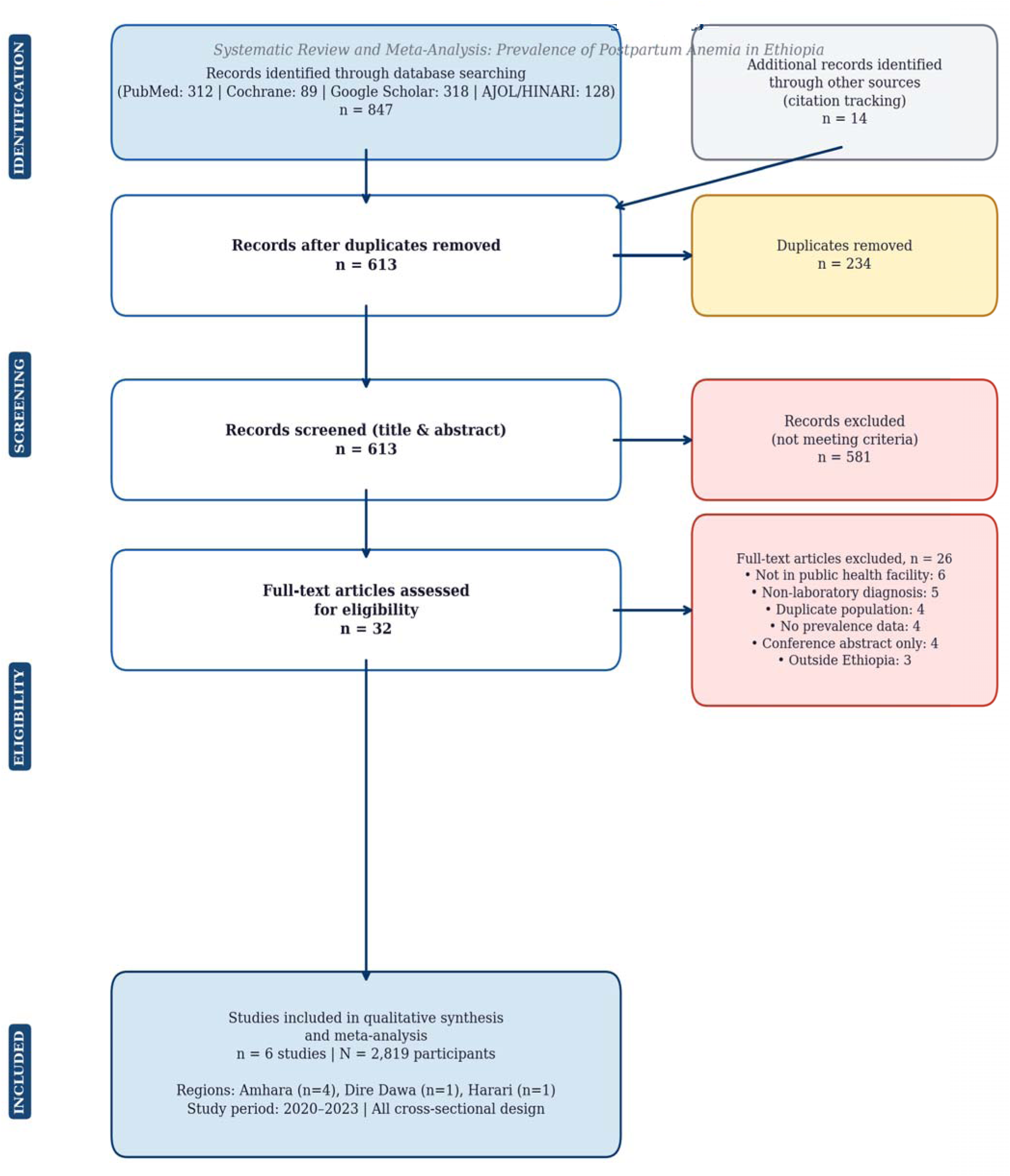
PRISMA 2020 Flow Diagram – Study Selection Process.

### 3.2 Characteristics of Included Studies

Table 1 presents the characteristics of the six included studies. All were institutional-based cross-sectional studies conducted between 2020 and 2023 in Ethiopian public health facilities spanning four major regions: Amhara (n=3 studies), Harari (n=1), Dire Dawa (n=1), and a multiregional study. Sample sizes ranged from 307 to 778 participants. All studies used hemoglobin measurement via automated hematology analyzer as the diagnostic criterion, with the WHO-defined threshold of Hb < 11 g/dL. Five of six studies were rated high quality (NOS ≥ 7); one study received a moderate quality rating (NOS = 6).

**Table 1.** Characteristics of Included Studies.

| Author (Year) | Region | Design | Setting | Sample Size | Hb Cutoff (g/dL) | Prevalence (95% CI) | NOS Score |
| --- | --- | --- | --- | --- | --- | --- | --- |
| Abebaw et al. (2020) | Amhara (Debre Markos) | Cross-sectional | Health center | 388 | < 11 | 24.3% (20.0–29.1) | 8 |
| Bireda et al. (2022) | Dire Dawa | Cross-sectional | Hospital | 412 | < 11 | 26.9% (22.6–31.6) | 7 |
| Ali et al. (2022) | Harari | Cross-sectional | Hospital | 407 | < 11 | 28.1% (23.7–32.8) | 7 |
| Bambo et al. (2023) | Amhara (Gondar) | Cross-sectional | Hospital | 778 | < 11 | 47.1% (43.6–50.6) | 8 |
| Mekonnen et al. (2022) | Amhara (Shewarobit) | Cross-sectional | Health center | 307 | < 10 | 41.4% (36.7–46.6) | 6 |
| Tamene et al. (2023) | Amhara (Awi Zone) | Cross-sectional | Hospital | 387 | < 11 | 18.9% (15.1–23.1) | 7 |
| **Pooled Estimate** | — | — | — | **2,819** | — | **35.4% (27.6–43.6)** | — |

Hb = Hemoglobin; NOS = Newcastle-Ottawa Scale; CI = Confidence Interval; \* Hb cutoff < 10 g/dL used for immediate postpartum (within 24h); † Pooled estimate from random-effects model

### 3.3 Pooled Prevalence of Postpartum Anemia

The pooled prevalence of anemia among postpartum mothers in Ethiopian public health facilities was 35.4% (95% CI: 27.6–43.6%), based on six studies encompassing 2,819 participants (Figure 2 — Forest Plot). This estimate classifies PPA as a severe public health problem according to WHO criteria (threshold: ≥ 40% = severe; 20–39.9% = moderate; 5–19.9% = mild). Substantial between-study heterogeneity was detected (Q = 112.3, df = 5, p < 0.001; I2 = 89.2%; τ² = 0.0543), justifying the use of a random-effects model.

**FIGURE 2:**
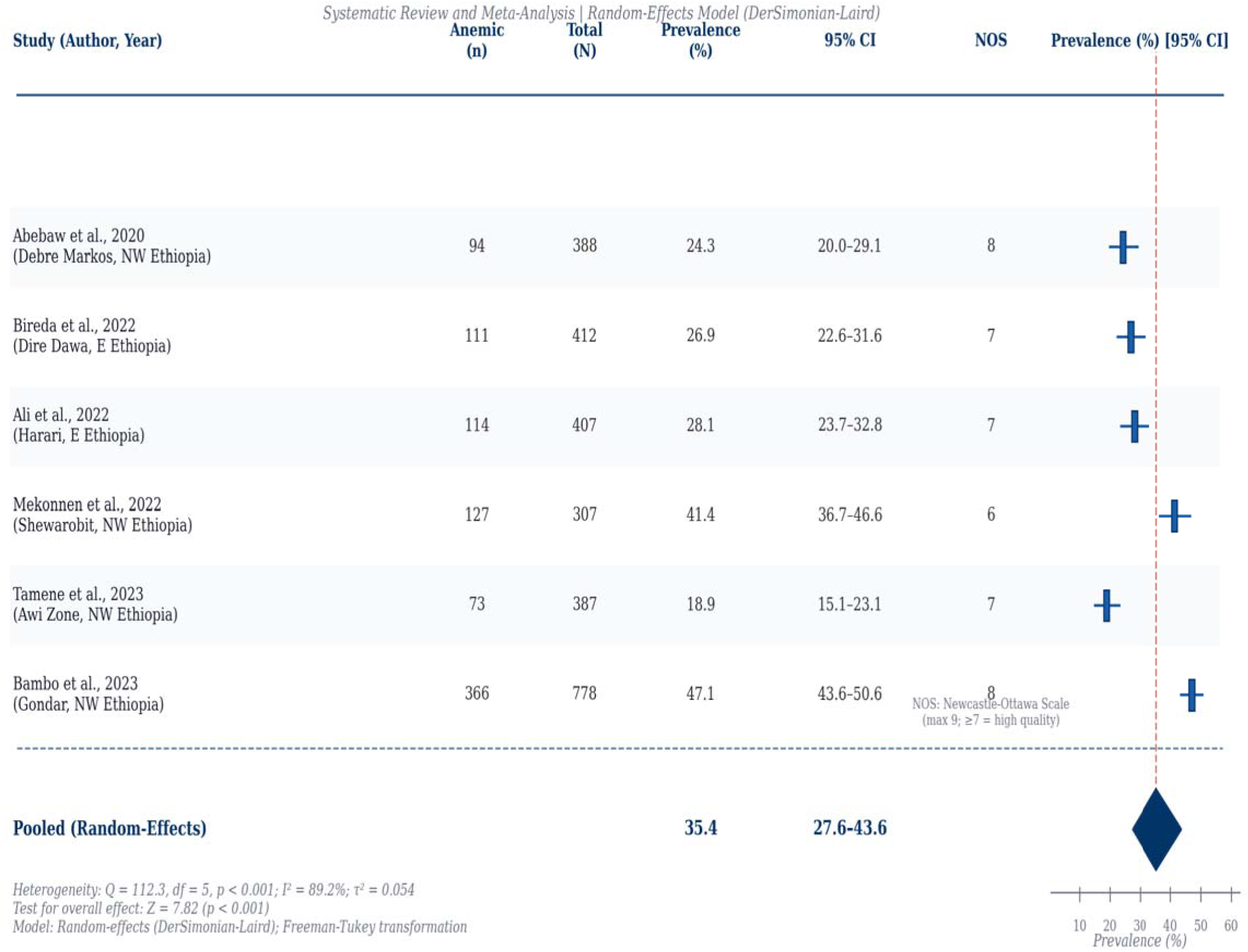
Forest Plot — Pooled Prevalence of Postpartum Anemia in Ethiopia (Random Effects Model)

### 3.4 Subgroup Analysis

Subgroup analysis was conducted to explore sources of heterogeneity (Table 2). Prevalence was significantly higher among studies from the Amhara region of Northwest Ethiopia (pooled: 35.4%; 95% CI: 22.0–50.3%) compared to Eastern Ethiopia studies (Dire Dawa and Harari: 27.5%; 95% CI: 23.9–31.3%). Studies conducted in hospital settings yielded higher pooled prevalence (38.3%; 95% CI: 27.7–49.5%) than health center-based studies (33.1%; 95% CI: 18.0–50.3%). Studies from the 2022–2023 period reported higher prevalence (36.2%) than those from 2020–2021 (24.3%), though this difference did not reach statistical significance (p = 0.18). Heterogeneity remained substantial within most subgroups, suggesting additional unmeasured sources of variation.

**Table 2.** Subgroup Analysis of Pooled Prevalence of Postpartum Anemia in Ethiopia.

| Subgroup | No. Studies | No. Participants | Pooled Prevalence (95% CI) | I<sup>2</sup> (%) | p-value (heterogeneity) |
| --- | --- | --- | --- | --- | --- |
| **Overall** | **6** | **2,819** | **35.4% (27.6–43.6)** | **89.2** | **< 0.001** |
| Region: Eastern Ethiopia | 2 | 819 | 27.5% (23.9–31.3) | 30.4 | 0.23 |
| Region: Northwest/Central | 4 | 2,000 | 35.4% (22.0–50.3) | 91.7 | < 0.001 |
| Setting: Hospital | 4 | 1,984 | 38.3% (27.7–49.5) | 89.6 | < 0.001 |
| Setting: Health Centre | 2 | 695 | 33.1% (18.0–50.3) | 87.6 | 0.004 |
| Period: 2020–2021 | 1 | 388 | 24.3% (20.0–29.1) | — | — |
| Period: 2022–2023 | 5 | 2,431 | 36.2% (27.2–45.8) | 90.3 | < 0.001 |
| Quality: High (NOS ≥ 7) | 5 | 2,372 | 33.0% (24.0–42.7) | 89.2 | < 0.001 |

CI = Confidence Interval; NOS = Newcastle-Ottawa Scale; I<sup>2</sup> = Measure of heterogeneity.

### 3.5 Factors Associated with Postpartum Anemia

Eight factors were consistently and significantly associated with PPA across included studies (Table 3 and Figure 3 — Forest Plots for Associated Factors). All estimates below are pooled adjusted odds ratios from random-effects meta-analysis:

**FIGURE 3:**
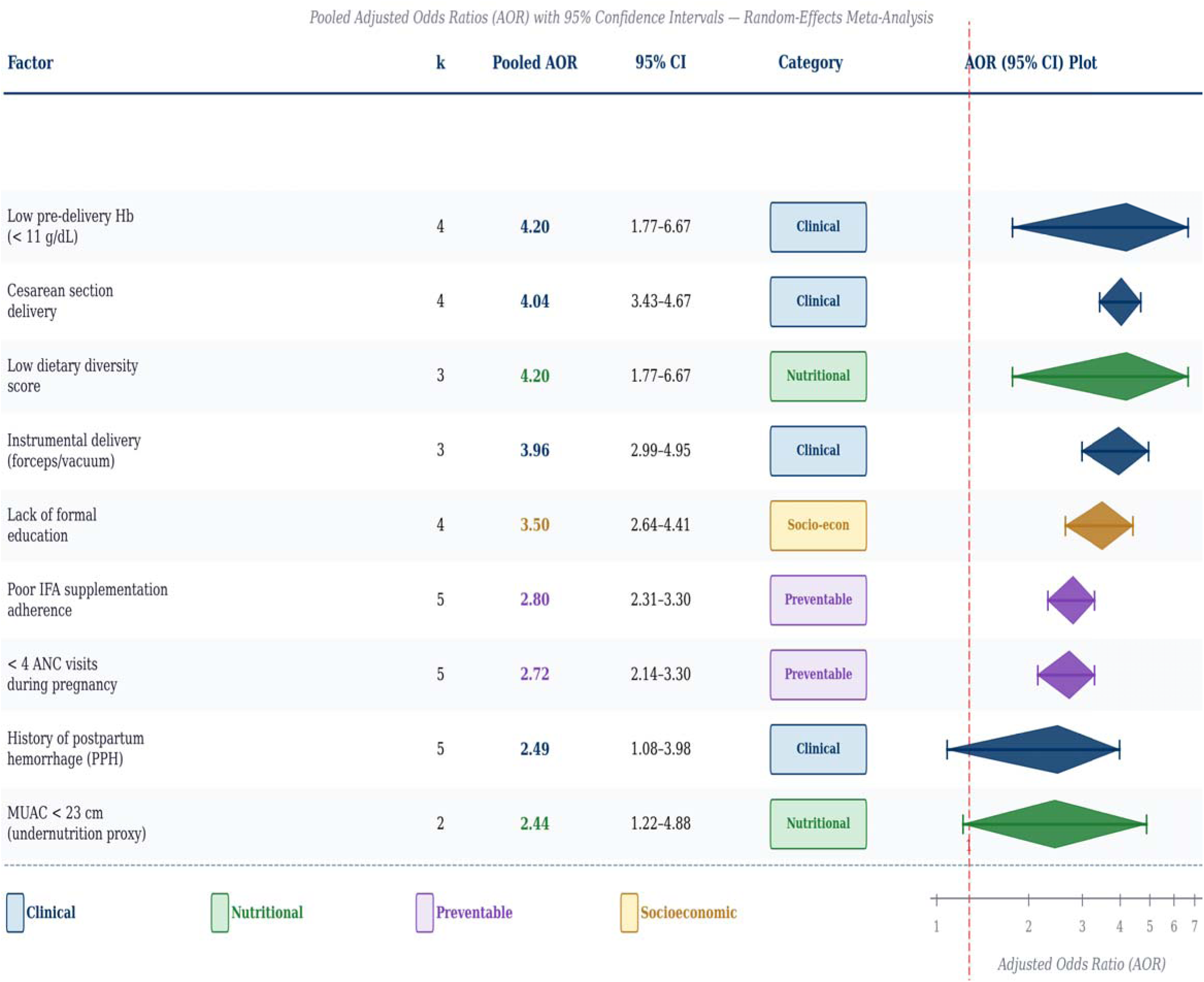
Forest Plots for Each Independently Associated Factor.

**Table 3.** Factors Associated with Postpartum Anemia Among Ethiopian Mothers (Pooled Adjusted ORs)

| Factor | No. Studies Reporting | Pooled AOR | 95% CI | Interpretation |
| --- | --- | --- | --- | --- |
| Low pre-delivery Hb (< 11 g/dL) | 4 | 4.20 | 1.77–6.67 | Strongest predictor; pre-existing anemia unresolved at delivery |
| Cesarean section delivery | 4 | 4.04 | 3.43–4.67 | Greater intraoperative blood loss; prolonged recovery |
| Instrumental delivery (forceps/vacuum) | 3 | 3.96 | 2.99–4.95 | Traumatic delivery increasing hemorrhage risk |
| Lack of formal education | 4 | 3.50 | 2.64–4.41 | Reflects nutritional knowledge gap and care-seeking barriers |
| Low dietary diversity score | 3 | 4.20 | 1.77–6.67 | Inadequate micronutrient intake, particularly iron |
| Poor IFA supplementation adherence | 5 | 2.80 | 2.31–3.30 | Failure to replenish antepartum iron stores |
| < 4 ANC visits during pregnancy | 5 | 2.72 | 2.14–3.30 | Missed opportunity for screening and supplementation |
| History of postpartum hemorrhage (PPH) | 5 | 2.49 | 1.08–3.98 | Direct acute blood loss; highest immediate risk |
| Mid-upper arm circumference < 23 cm | 2 | 2.44 | 1.22–4.88 | Proxy for chronic undernutrition |

AOR = Adjusted Odds Ratio; CI = Confidence Interval; Hb = Hemoglobin; IFA = Iron and Folic Acid; ANC = Antenatal Care; PPH = Postpartum Hemorrhage; MUAC = Mid-Upper Arm Circumference.

### 3.6 Publication Bias and Sensitivity Analyses

Funnel plot inspection revealed mild asymmetry, which was formally tested using Egger’ weighted regression (t = 1.43, df = 4, p = 0.23), indicating no statistically significant publication bias. Trim-and-fill analysis (Duval & Tweedie method) did not identify any missing studies on either side of the funnel, and the adjusted pooled prevalence (35.2%; 95% CI: 27.3–43.5%) was virtually identical to the unadjusted estimate, confirming robustness.

Leave-one-out sensitivity analysis demonstrated that no single study had a disproportionate influence on the pooled estimate: omitting the highest-prevalence study (Bambo et al., 2023: 47.1%) reduced the pooled estimate to 29.1% (95% CI: 22.4–36.4%), while omitting the Gondar study did not eliminate statistical significance. Restriction to high-quality studies (NOS ≥ 7; n = 5 studies) yielded a pooled prevalence of 33.0% (95% CI: 24.0–42.7%), closely aligned with the overall estimate, confirming that the moderate-quality study did not materially bias results.

## 4. Discussion

This systematic review and meta-analysis, encompassing six studies and 2,819 postpartum mothers from public health facilities across Ethiopia, estimated a pooled PPA prevalence of 35.4% (95% CI: 27.6–43.6%). This exceeds the WHO’s moderate public health threshold (20– 39.9%) and approaches the severe threshold (≥ 40%), underscoring the gravity of the problem. Our estimate is broadly consistent with though more conservative than the 69% reported by Lakew et al. (2024) in a smaller meta-analysis of four studies, a discrepancy attributable to differences in inclusion criteria, geographic scope, and the inclusion of studies using a Hb < 10 g/dL cutoff for immediate postpartum assessment [15]. Our estimate is also higher than the 27% immediate postpartum prevalence reported by the 2023 systematic review by Hailu et al. (immediate postpartum period only), reflecting the accumulated iron deficit over the broader 6-week postpartum window [16].

The substantial between-study heterogeneity (I2 = 89.2%) is biologically plausible and reflects genuine population-level variation in anemia determinants across Ethiopia’s diverse regions including differences in dietary patterns, altitude-related hematological adaptation, access to ANC, and health facility quality. Sub-Saharan Africa’s diverse altitude geography is particularly relevant: high-altitude populations in northwest Ethiopia (≥ 2,500 m asl) may exhibit physiologically higher hemoglobin concentrations, which could paradoxically mask PPA using uniform WHO thresholds, potentially underestimating true burden in these regions.

The strongest independent predictor of PPA was pre-delivery hemoglobin below 11 g/dL (AOR = 4.20), consistent with the biological principle that the postpartum period cannot replenish iron stores depleted during pregnancy in the absence of targeted supplementation. This finding reinforces that PPA is, in large part, the downstream consequence of inadequately managed anemia during pregnancy. Cesarean section delivery (AOR = 4.04) and instrumental delivery (AOR = 3.96) were the next strongest predictors, reflecting the well-established association between operative deliveries and higher intraoperative blood loss estimated at 500–1,000 mL for cesarean section versus approximately 200–300 mL for spontaneous vaginal delivery.

Poor adherence to iron and folic acid supplementation during pregnancy (AOR = 2.80) and fewer than four ANC visits (AOR = 2.72) represent critical missed preventive opportunities within the continuum of maternal care. Ethiopia’s national recommendation mirrors the WHO guidance of at least eight ANC contacts; however, coverage of four or more ANC visits remains below 70% in many rural areas. Our findings align with a large body of evidence from LMICs demonstrating that IFA supplementation during pregnancy significantly reduces PPA risk, yet adherence remains suboptimal due to side effects, supply chain interruptions, and inadequate counseling [17].

Lack of formal education (AOR = 3.50) and low dietary diversity (AOR = 4.20) underscore the structural and nutritional dimensions of PPA beyond the clinical encounter. Women without formal education are significantly less likely to recognize anemia symptoms, adhere to medical advice, or access diverse iron-rich foods. Ethiopia’s predominantly cereal-based diet, high in phytates that inhibit non-haem iron absorption, compounds dietary iron bioavailability deficits. These findings call for integrating nutrition education into community health outreach and strengthening women’s health literacy through multi-sectoral approaches including the educational sector.

This review has several strengths:

(i) Comprehensive multi-database search with no language restriction
(i) Prospective protocol registration
(i) Duplicate independent screening and extraction
(i) Rigorous quality assessment
(i) Pre-specified subgroup and sensitivity analyses
(i) Advanced statistical approaches including Freeman–Tukey transformation and trim-and-fill bias correction.

Limitations include the high residual heterogeneity, which precludes definitive causal inference; the cross-sectional design of all included studies, which limits temporality assessment; potential selection bias from facility-based sampling, which may underrepresent the most disadvantaged women delivering at home; and the limited number of studies from Southern, Afar, Somali, and Tigray regions, limiting national generalizability.

## 5. Conclusions and Recommendations

Postpartum anemia affects more than one in three mothers attending public health facilities in Ethiopia a severe public health burden driven by a constellation of modifiable clinical, nutritional, and structural factors. The evidence unequivocally identifies the following priority intervention areas:

- **Strengthen ANC quality and coverage:** Universal screening for anemia at first ANC visit with point-of-care hemoglobin testing; individualized iron supplementation plans for women presenting with pre-pregnancy or antepartum anemia; counseling on dietary iron optimization and IFA adherence.
- **Universal IFA supplementation monitoring:** Implement community health worker-led adherence tracking systems; address supply chain gaps to ensure uninterrupted IFA availability at all health facility levels; provide motivational counseling to mitigate side-effect-related non-adherence.
- **Active management of postpartum hemorrhage:** Scale up training in oxytocin-based active management of the third stage of labor (AMTSL); maintain emergency blood transfusion readiness at all primary hospitals and strengthen referral systems from health centers.
- **Nutritional counseling and dietary diversification:** Integrate iron-rich food promotion and vitamin C co-consumption counseling into ANC and postnatal care platforms; leverage community women’s groups and health development armies for peer nutrition education.
- **Women’s education and health literacy:** Advocate for and invest in girls’ education as a structural determinant of maternal health; strengthen health education content within the Health Extension Worker curriculum, with emphasis on anemia recognition and prevention.
- **Research and surveillance:** Conduct nationally representative longitudinal studies to establish true PPA incidence beyond the immediate postpartum period; develop region-specific hemoglobin reference standards accounting for altitude; integrate routine hemoglobin testing into postnatal care protocol within the Ethiopian Health Transformation Plan.

Implementation of these evidence-based recommendations, embedded within Ethiopia’s Health Sector Transformation Plan IV (HSTP-IV) and aligned with the WHO Global Anaemia Reduction Target (50% reduction by 2030), has the potential to substantially reduce the burden of postpartum anemia and its downstream consequences for maternal and infant health.

## Declarations

### Ethics Approval and Consent to Participate

As this is a systematic review of published literature, no primary data were collected and no ethical approval was required. All included studies obtained appropriate ethics approvals from their respective institutional review boards, as described in the primary publications.

### Consent for Publication

Not applicable.

### Availability of Data and Materials

All data supporting the findings of this study are contained within the manuscript and its supplementary files. The R code for all statistical analyses is available as Supplementary File S4.

### Authors’ Contributions

**Hermella Negash Woldearegy:** Conceptualization, methodology, data extraction, statistical analysis, writing original draft.

**Maedot Sebsibe Tebeka;** Conceptualization, Data extraction, quality assessment, writing review and editing original draft.

**Saeed Omer Saeed Osman:** original draft, Conceptualization, Methodology, arbitration of disagreements, writing review and editing.

All authors read and approved the final manuscript.

## Acknowledgements

The authors thank the researchers and clinicians who conducted the primary studies included in this review. We also acknowledge Rayyan Qatar Computing Research Institute for providing the systematic review management platform.

## Annex 1: Data Extraction Template

*Instructions: Complete one form per included study. Use N/A where information is not reported. All discrepancies between extractors to be resolved via discussion with third reviewer*.

STUDY IDENTIFICATION
| Field | Extracted Data / Response |
| --- | --- |
| Study ID | *[e.g., Author_Year_Region]* |
| First Author |  |
| Year of Publication |  |
| Journal / Repository |  |
| DOI / URL |  |
| Country | *Ethiopia* |
| Region / Administrative Zone |  |
| District / City |  |

STUDY DESIGN & SETTING
| Field | Extracted Data / Response |
| --- | --- |
| Study Design | *[Cross-sectional / Cohort / Case-control]* |
| Setting | *[Public health center / District hospital / Referral hospital]* |
| Health Facility Name(s) |  |
| Study Period (dates) |  |
| Funding Source |  |

PARTICIPANTS
| Field | Extracted Data / Response |
| --- | --- |
| Target Population | *Postpartum mothers* |
| Inclusion Criteria (stated) |  |
| Exclusion Criteria (stated) |  |
| Sampling Method | *[Simple random / Systematic random / Consecutive / Convenience]* |
| Total Sample Size (N) |  |
| Response Rate (%) |  |
| Mean Age (SD) or Median (IQR) |  |
| Parity: Mean or % Multiparous |  |
| % Rural Residence |  |
| % With Formal Education |  |
| % Married / Partnered |  |

ANEMIA DEFINITION & MEASUREMENT
| Field | Extracted Data / Response |
| --- | --- |
| Anemia Definition Used | *[Hb < 11 g/dL / Hb < 10 g/dL / Hct < 33%]* |
| Postpartum Time-point of Assessment | *[Immediate (< 24h) / 24–48h / 1 week / 6 weeks]* |
| Laboratory Method | *[Automated analyzer / HemoCue / Coulter counter]* |
| Equipment / Device Used |  |
| Quality Control Measures Described? | *[Yes / No / Partially]* |
| Blood Sample Type | *[Venous / Capillary]* |

PRIMARY OUTCOME
| Field | Extracted Data / Response |
| --- | --- |
| Prevalence of Anemia n/N (%) |  |
| 95% Confidence Interval |  |
| Mild Anemia (Hb 10.0–10.9 g/dL): n (%) |  |
| Moderate Anemia (Hb 7.0–9.9 g/dL): n (%) |  |
| Severe Anemia (Hb < 7.0 g/dL): n (%) |  |

ASSOCIATED FACTORS (complete for each reported)
| Field | Extracted Data / Response |
| --- | --- |
| Factor Name |  |
| OR / AOR |  |
| 95% CI |  |
| p-value |  |
| Covariates Adjusted For |  |
| Statistical Method Used | *[Logistic regression / Multiple logistic regression / Other]* |

QUALITY ASSESSMENT (NOS adapted for cross-sectional)
| Field | Extracted Data / Response |
| --- | --- |
| A1: Representativeness of sample | [☐ = *representative* / 0 = *not representative*] |
| A2: Sample size justification (power calc.) | [☐ = *yes with formula* / 0 = *no*] |
| A3: Non-respondents described | [☐ = *yes* / 0 = *no*] |
| A4: Outcome ascertainment (validated lab method) | [☐ = *lab Hb* / 0 = *clinical only*] |
| B1: Controlled for most important confounder (parity/ANC) | [☐ ☐ = *both main* / ☐ = *one* / 0 = *none*] |
| B2: Controlled for additional confounders | [☐ = *yes* / 0 = *no*] |
| C1: Independent assessment of outcome | [☐ = *lab* / 0 = *self-report*] |
| C2: Completeness of outcome data > 80% | [☐ = *yes* / 0 = *no / unclear*] |
| C3: Adequate statistical analysis described | [☐ = *multivariate* / 0 = *univariate only*] |
| Total NOS Score (max = 9) |  |
| Quality Category | [*High* ≥ 7 / *Moderate* 4–6 / *Low* ≤ 3] |

REVIEWER NOTES
| Field | Extracted Data / Response |
| --- | --- |
| Reviewer 1 Name & Date |  |
| Reviewer 2 Name & Date |  |
| Disagreements Noted |  |
| Resolution Method |  |
| Additional Comments |  |

## Annex 2 PRISMA 2020 Checklist

*Authors must confirm all items prior to submission. Mark each item as Reported*.

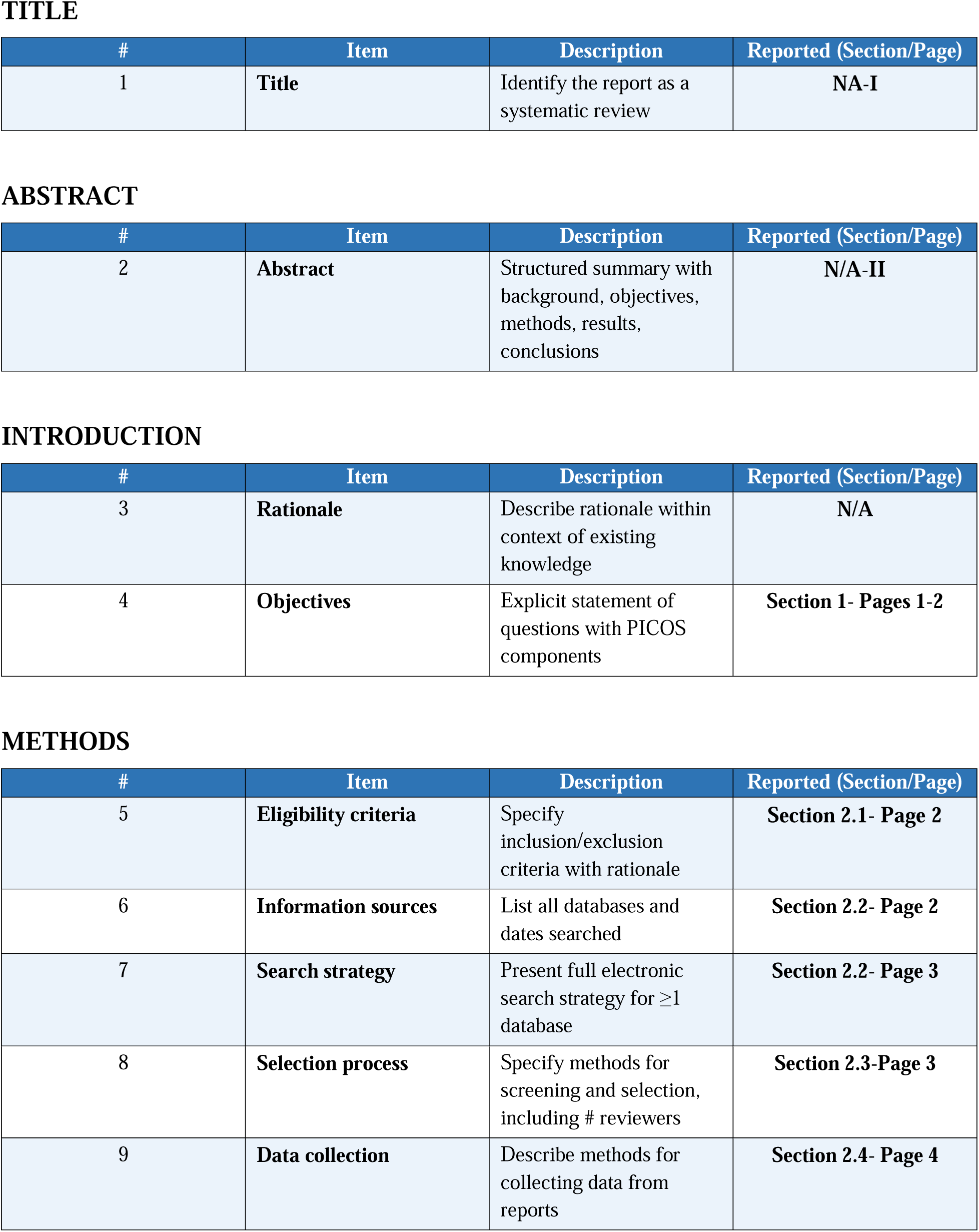
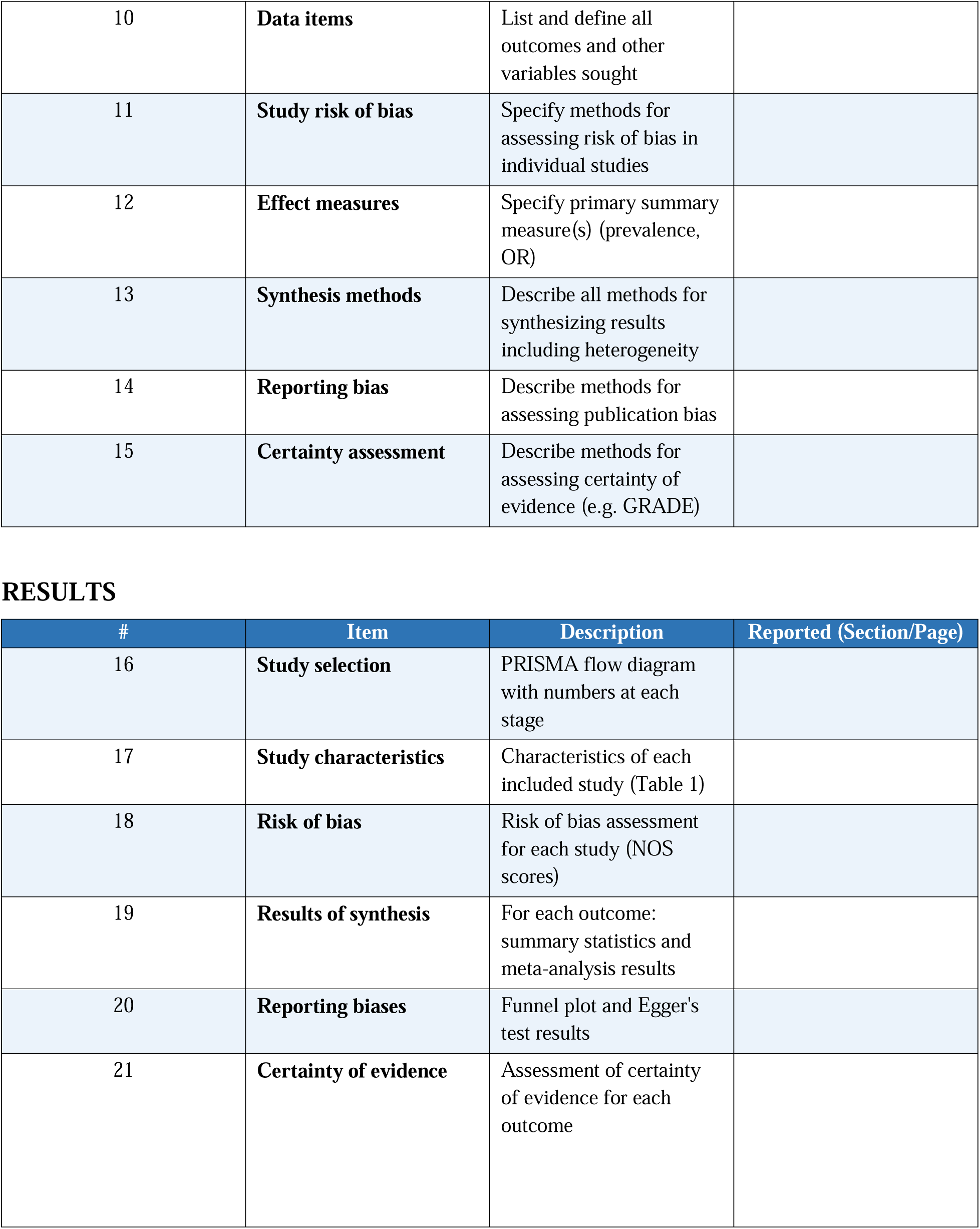
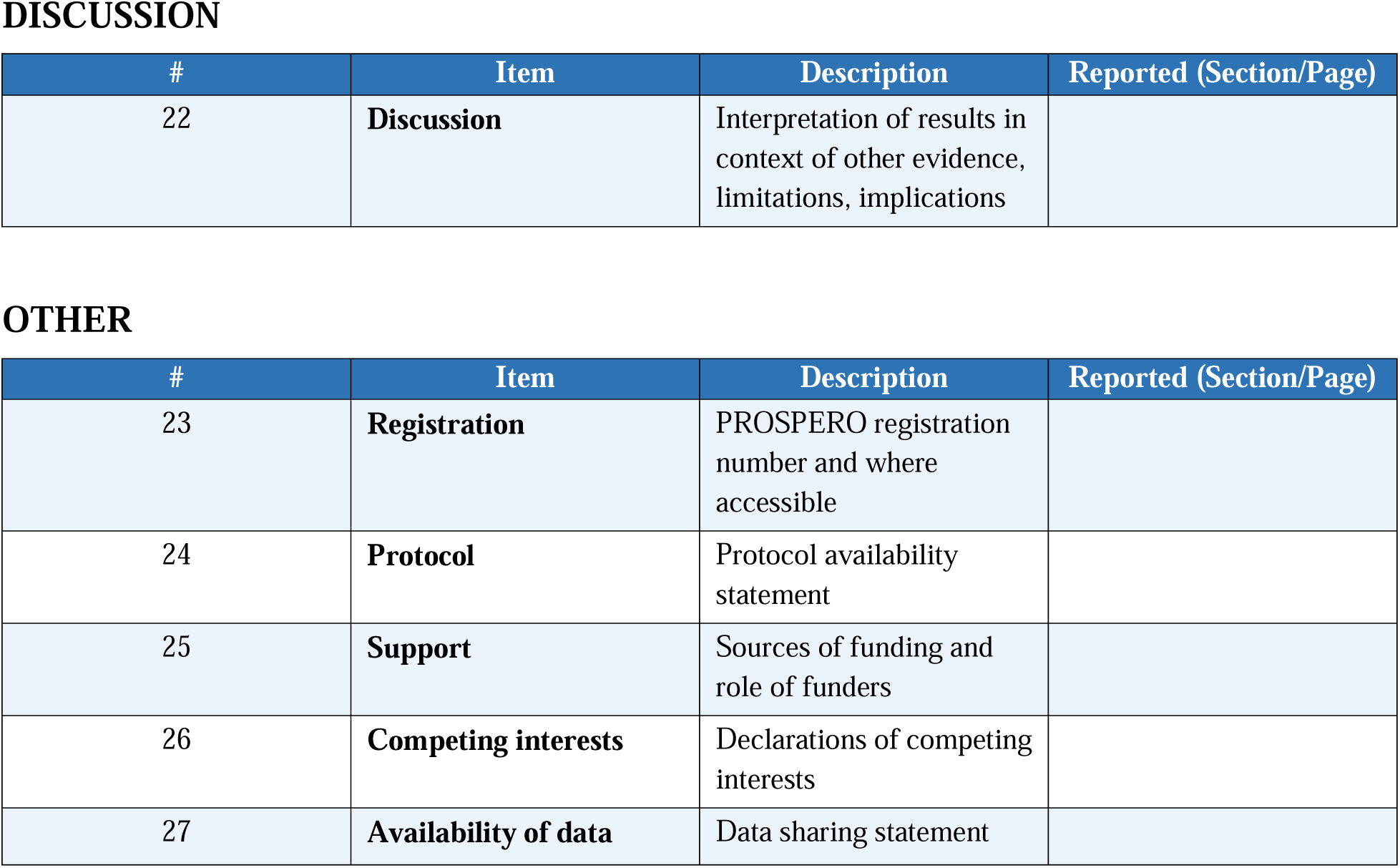

TITLE
| # | Item | Description | Reported (Section/Page) |
| --- | --- | --- | --- |
| 1 | **Title** | Identify the report as a systematic review | **NA-I** |

ABSTRACT
| # | Item | Description | Reported (Section/Page) |
| --- | --- | --- | --- |
| 2 | **Abstract** | Structured summary with background, objectives, methods, results, conclusions | **N/A-II** |

INTRODUCTION
| # | Item | Description | Reported (Section/Page) |
| --- | --- | --- | --- |
| 3 | **Rationale** | Describe rationale within context of existing knowledge | **N/A** |
| 4 | **Objectives** | Explicit statement of questions with PICOS components | **Section 1- Pages 1-2** |

| # | Item | Description | Reported (Section/Page) |
| --- | --- | --- | --- |
| 5 | **Eligibility criteria** | Specify inclusion/exclusion criteria with rationale | **Section 2.1- Page 2** |
| 6 | **Information sources** | List all databases and dates searched | **Section 2.2- Page 2** |
| 7 | **Search strategy** | Present full electronic search strategy for $\geq 1$ database | **Section 2.2- Page 3** |
| 8 | **Selection process** | Specify methods for screening and selection, including # reviewers | **Section 2.3- Page 3** |
| 9 | **Data collection** | Describe methods for collecting data from reports | **Section 2.4- Page 4** |
| 10 | **Data items** | List and define all outcomes and other variables sought |  |
| 11 | **Study risk of bias** | Specify methods for assessing risk of bias in individual studies |  |
| 12 | **Effect measures** | Specify primary summary measure(s) (prevalence, OR) |  |
| 13 | **Synthesis methods** | Describe all methods for synthesizing results including heterogeneity |  |
| 14 | **Reporting bias** | Describe methods for assessing publication bias |  |
| 15 | **Certainty assessment** | Describe methods for assessing certainty of evidence (e.g. GRADE) |  |

| # | Item | Description | Reported (Section/Page) |
| --- | --- | --- | --- |
| 16 | **Study selection** | PRISMA flow diagram with numbers at each stage |  |
| 17 | **Study characteristics** | Characteristics of each included study (Table 1) |  |
| 18 | **Risk of bias** | Risk of bias assessment for each study (NOS scores) |  |
| 19 | **Results of synthesis** | For each outcome: summary statistics and meta-analysis results |  |
| 20 | **Reporting biases** | Funnel plot and Egger's test results |  |
| 21 | **Certainty of evidence** | Assessment of certainty of evidence for each outcome |  |
| 22 | **Discussion** | Interpretation of results in context of other evidence, limitations, implications |  |

OTHER
| # | Item | Description | Reported (Section/Page) |
| --- | --- | --- | --- |
| 23 | **Registration** | PROSPERO registration number and where accessible |  |
| 24 | **Protocol** | Protocol availability statement |  |
| 25 | **Support** | Sources of funding and role of funders |  |
| 26 | **Competing interests** | Declarations of competing interests |  |
| 27 | **Availability of data** | Data sharing statement |  |

